# Interpretable machine learning model for predicting kidney failure among CAKUT children in multicenter large-scale study

**DOI:** 10.64898/2026.02.08.26345871

**Authors:** Tianyi Liu, Helin Wang, Jialu Liu, Xinyuan Zhao, Yilan Xia, Xiaowen Wang, Yulin Kang, Cuihua Liu, Xiaojie Gao, Xiaoyun Jiang, Jianhua Mao, Yufeng Li, Aihua Zhang, Mo Wang, Haitao Bai, Tong Shen, Xiqiang Dang, Dexuan Wang, Ruifeng Zhang, Yihan Lu, Qian Shen, Sheng Nie, Yang Chen, Hong Xu, Yihui Zhai, the CCGKDD and CRDS study investigators

## Abstract

Congenital anomalies of the kidney and urinary tract (CAKUT) are the leading cause of pediatric kidney failure, but predicting individual progression remains challenging. This multicenter study developed and validated POCC, a machine learning model for predicting kidney failure risk at 1, 3, and 5 years post-diagnosis in CAKUT patients. Two versions were created using data from 2,249 children. The general model achieved internal AUCs of 0.93–0.99 and external AUCs of 0.90–0.98 and 0.81– 0.90 in two independent validations at pediatric and general hospitals, respectively. The specialized model, integrating congenital-hereditary features, achieved internal AUCs of 0.93–0.99 and external AUCs of 0.91–0.96 in pediatric hospitals. Deployed online, POCC demonstrated 90.7% accuracy in real-world validation, with the specialized model reaching 100% sensitivity and specificity for 5-year predictions. As the first tool for multi-timepoint risk prediction across diverse CAKUT subphenotypes per patient, POCC has strong potential to support personalized management.

## Introduction

Congenital anomalies of the kidney and urinary tract (CAKUT) constitute a heterogeneous group of disorders characterized by structural malformations of the urinary system and represent one of the most common birth defects, accounting for approximately 20–50% of congenital anomalies^1^. CAKUT is the predominant cause of pediatric chronic kidney disease (CKD) and kidney failure and is responsible for 50–70% of children who require dialysis or kidney transplantation^2^. Moreover, CAKUT is associated with elevated risks of developing hypertension and adult CKD. Among patients who progress to kidney failure before age 30, approximately 40% have underlying CAKUT^3^. Given its diverse clinical phenotypes, CAKUT has a broad spectrum of prognoses, ranging from mild cases with preserved renal function to life-threatening cases that rapidly progress to kidney failure. Among patients with CAKUT who progress to kidney failure, approximately 20–25% develop kidney injury before 18 years of age. A 20-year registry study by the European Renal Association and European Dialysis and Transplant Association that included more than 200,000 patients with kidney failure reported that approximately one-third of those with CAKUT-related kidney failure initiated renal replacement therapy before age 20^4^. These findings highlight the need for early and precise detection of high-risk individuals to improve outcomes and guide personalized, cost-effective clinical management and long-term care strategies.

Several population-level risk factors for CAKUT prognosis have been identified, including CAKUT subphenotype, CKD stage at baseline, proteinuria, hypertension, anemia, short stature, preterm birth, low birth weight, and small for gestational age^5–8^. However, in clinical practice, patients frequently present with complex combinations of interacting risk factors, and even within the same CAKUT subphenotype, the rate of renal function decline can vary considerably. For example, the incidence of chronic kidney injury among children with a functional solitary kidney ranges from 0% to 45% over a 7–10 year follow-up period^9,10^. The increasing availability of clinical data and advances in computational methods have accelerated the development of data-driven modeling approaches. However, traditional statistical models remain limited, as they often rely on population-level characteristics and incorporate limited variables. Consequently, these models frequently fail to provide individualized, precise risk predictions^11,12^. As a branch of artificial intelligence, machine learning (ML) offers an effective alternative^13^ that overcomes the limitations of conventional linear models, such as the reliance on manual variable selection and challenges in handling nonlinear associations. However, clinical ML models for CAKUT remain limited. The few published studies have been restricted to a single subphenotype and were limited by single-center designs, small sample sizes, composite endpoints, reliance on a single algorithm, and lack of interpretability^14,15^. To date, no prognostic model has been developed for predicting outcomes across multiple CAKUT subphenotypes in children.

In this study, we developed a suite of ML-based models (general and specialized versions) to predict kidney failure outcomes in CAKUT children at 1, 3, and 5 years after initial diagnosis using baseline clinical, renal, and extrarenal phenotypes, and genetic features. We compared the performance of these models and validated them on two independent, external, multicenter nationwide cohorts to assess generalizability. We then employed the Shapley Additive exPlanations (SHAP) algorithm to identify and elucidate core features in model decision-making. Finally, we established a user-friendly online platform and conducted real-world validation in authentic clinical environments to evaluate the potential for translation. Figure 1 illustrates the overall design of the framework.

**Fig. 1:**
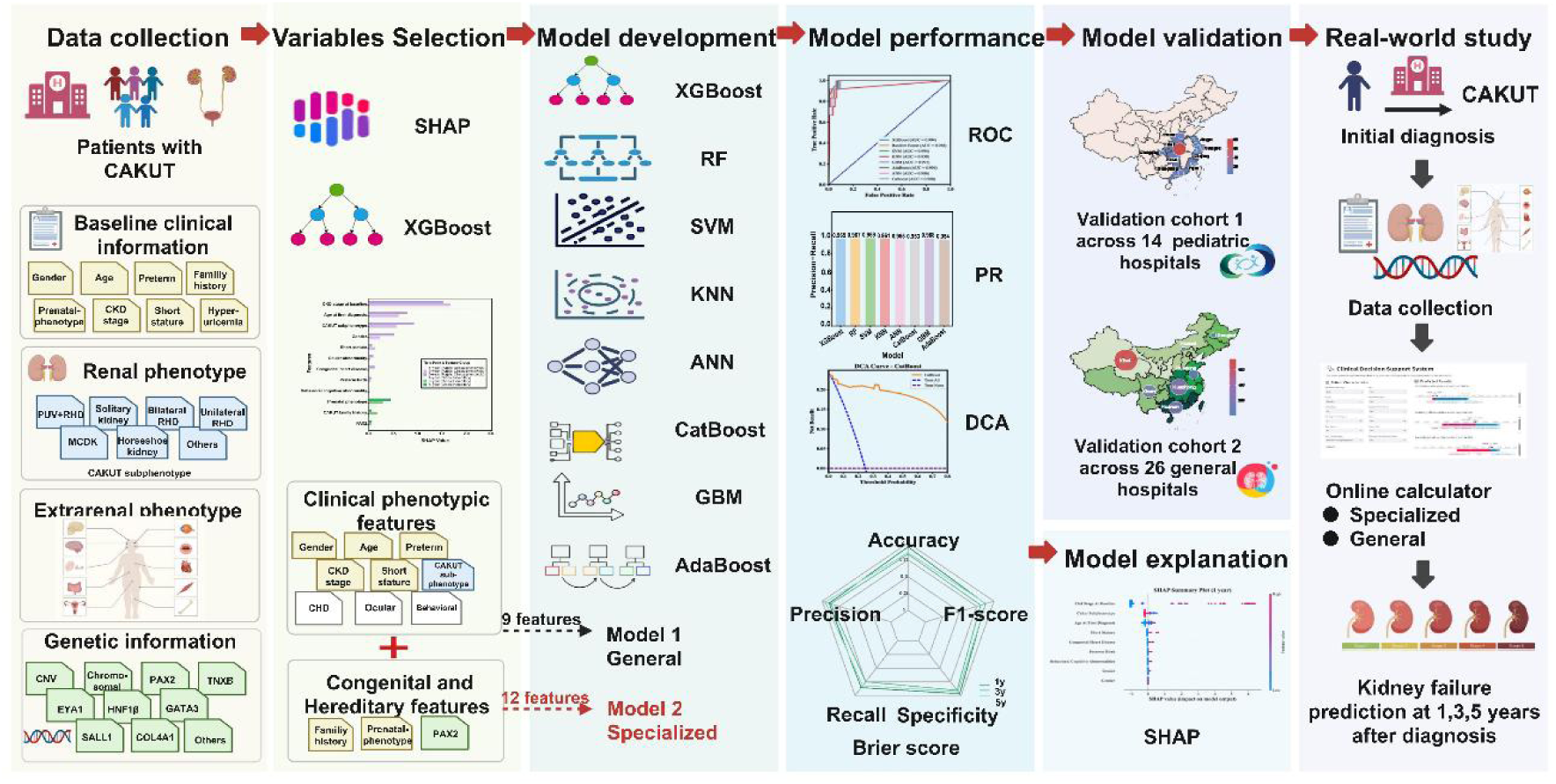
Overall design of the framework. CAKUT, congenital anomalies of the kidney and urinary tract. CKD, chronic kidney disease; PUV, posterior urethral valves; RHD, renal hypodysplasia; MCDK, multicystic dysplastic kidney; CNV, copy number variation; XGBoost, extreme gradient boosting; SHAP, SHapley Additive exPlanations; CHD, congenital heart disease; SVM, support vector machine; ANN, artificial neural network; KNN, k-nearest neighbors; RF, random forest; GBM, gradient boosting machine; AdaBoost, adaptive boosting; CatBoost, categorical boosting; ROC, receiver operating characteristic curve; PR, precision recall; DCA, decision curve analysis.

## Results

### Patient characteristics

Baseline characteristics of patients in the four cohorts are detailed in Table 1. A total of 2,249 patients were included in the analysis. The incidence of kidney failure in the derivation cohort (n=570), validation cohort 1 (n=142), validation cohort 2 (n=1,483), and real-world cohort (n=54) was 15.6%, 24.6%, 14.9%, and 24.1%, respectively. Patients in the derivation cohort (median age 0.62 years) and real-world cohort (median age 0.25 years) were significantly younger than those in the two external validation cohorts (3.15 and 5.63 years, respectively). Males accounted for more than 60% of participants across all cohorts.

**Table 1:**
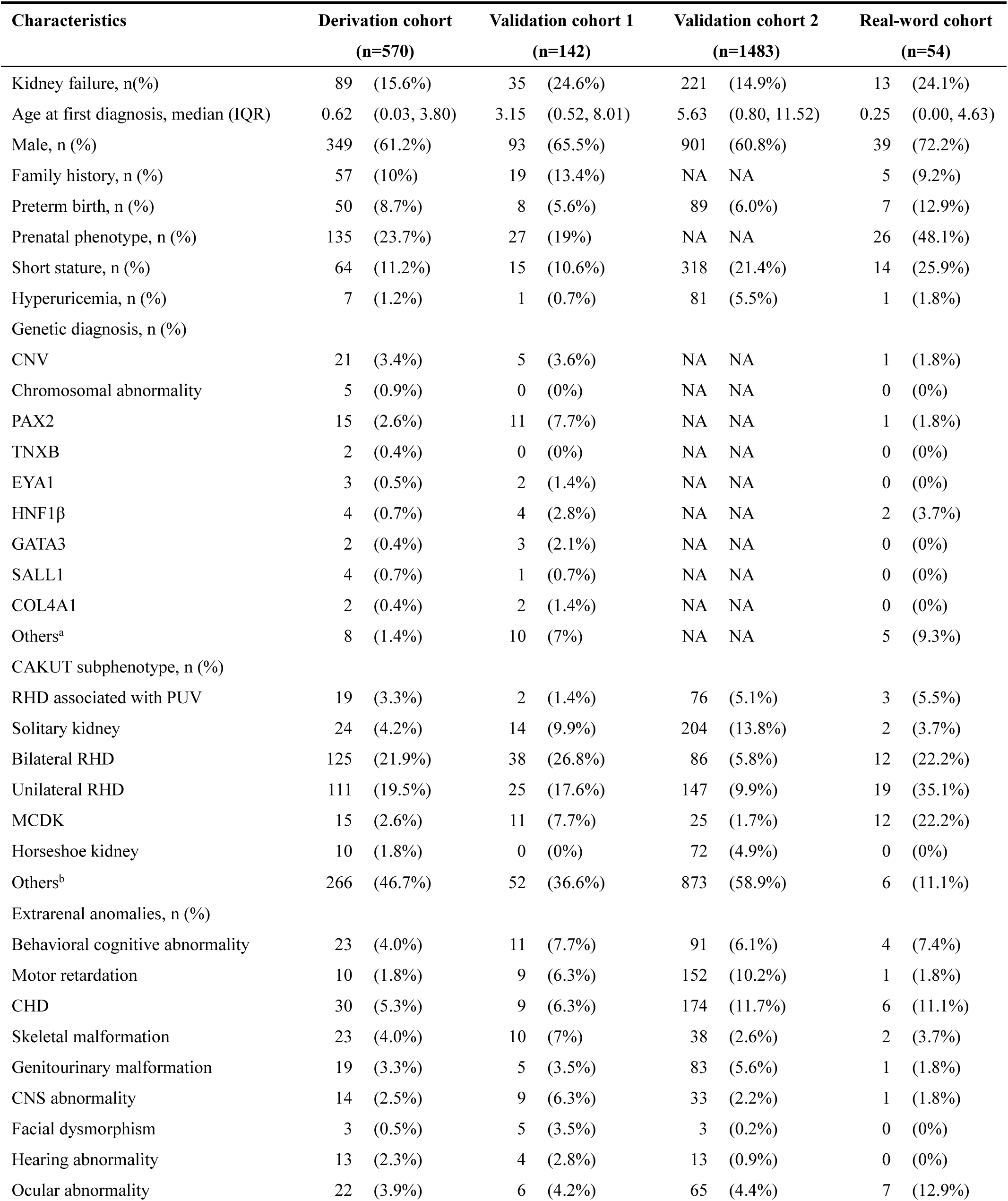
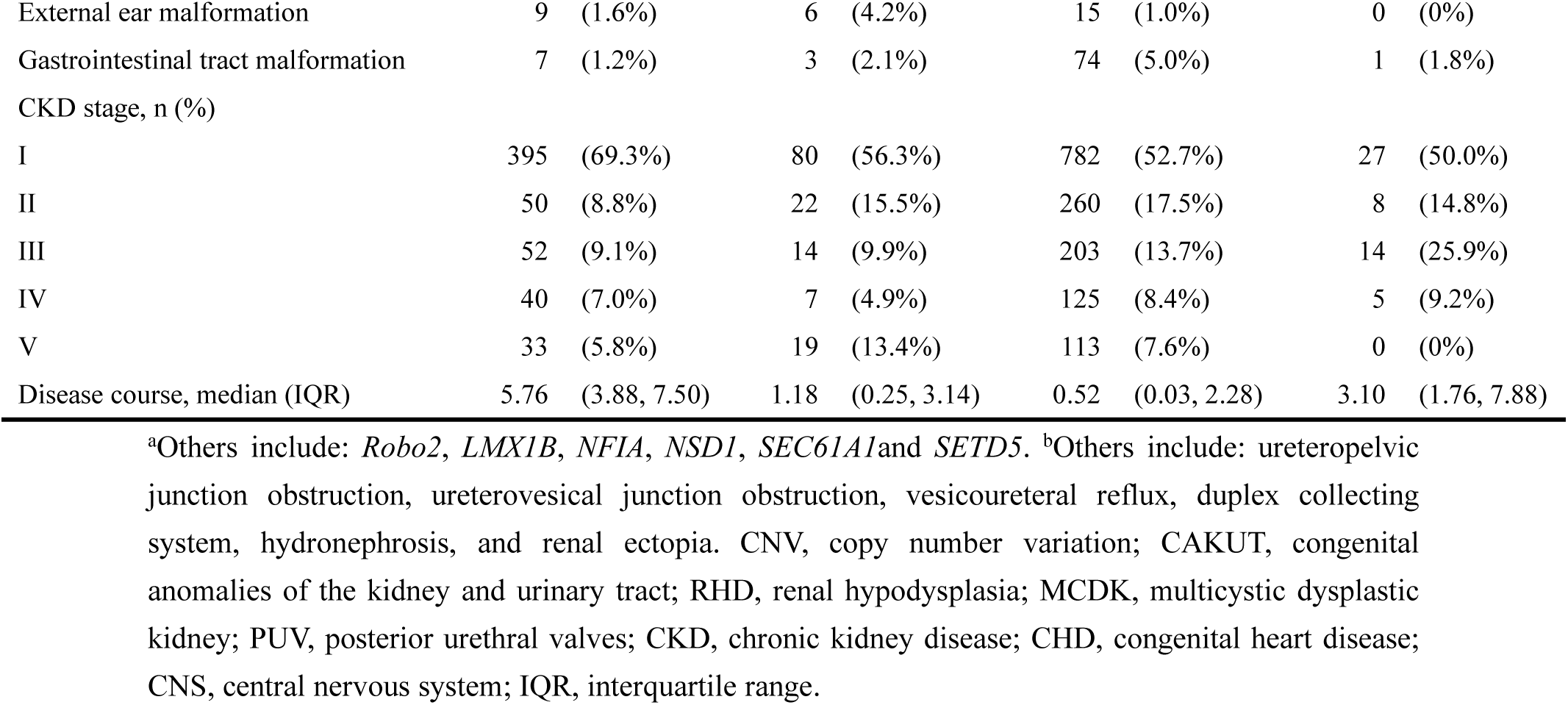
Baseline characteristics of patients with CAKUT in the derivation cohort and validation cohorts at the time of diagnosis.

| Characteristics | Derivation cohort<br>(n=570) |  | Validation cohort 1<br>(n=142) |  | Validation cohort 2<br>(n=1483) |  | Real-word cohort<br>(n=54) |  |
| --- | --- | --- | --- | --- | --- | --- | --- | --- |
| Kidney failure, n(%) | 89 | (15.6%) | 35 | (24.6%) | 221 | (14.9%) | 13 | (24.1%) |
| Age at first diagnosis, median (IQR) | 0.62 | (0.03, 3.80) | 3.15 | (0.52, 8.01) | 5.63 | (0.80, 11.52) | 0.25 | (0.00, 4.63) |
| Male, n (%) | 349 | (61.2%) | 93 | (65.5%) | 901 | (60.8%) | 39 | (72.2%) |
| Family history, n (%) | 57 | (10%) | 19 | (13.4%) | NA | NA | 5 | (9.2%) |
| Preterm birth, n (%) | 50 | (8.7%) | 8 | (5.6%) | 89 | (6.0%) | 7 | (12.9%) |
| Prenatal phenotype, n (%) | 135 | (23.7%) | 27 | (19%) | NA | NA | 26 | (48.1%) |
| Short stature, n (%) | 64 | (11.2%) | 15 | (10.6%) | 318 | (21.4%) | 14 | (25.9%) |
| Hyperuricemia, n (%) | 7 | (1.2%) | 1 | (0.7%) | 81 | (5.5%) | 1 | (1.8%) |
| Genetic diagnosis, n (%) |  |  |  |  |  |  |  |  |
| CNV | 21 | (3.4%) | 5 | (3.6%) | NA | NA | 1 | (1.8%) |
| Chromosomal abnormality | 5 | (0.9%) | 0 | (0%) | NA | NA | 0 | (0%) |
| PAX2 | 15 | (2.6%) | 11 | (7.7%) | NA | NA | 1 | (1.8%) |
| TNXB | 2 | (0.4%) | 0 | (0%) | NA | NA | 0 | (0%) |
| EYA1 | 3 | (0.5%) | 2 | (1.4%) | NA | NA | 0 | (0%) |
| HNF1β | 4 | (0.7%) | 4 | (2.8%) | NA | NA | 2 | (3.7%) |
| GATA3 | 2 | (0.4%) | 3 | (2.1%) | NA | NA | 0 | (0%) |
| SALL1 | 4 | (0.7%) | 1 | (0.7%) | NA | NA | 0 | (0%) |
| COL4A1 | 2 | (0.4%) | 2 | (1.4%) | NA | NA | 0 | (0%) |
| Others<sup>a</sup> | 8 | (1.4%) | 10 | (7%) | NA | NA | 5 | (9.3%) |
| CAKUT subphenotype, n (%) |  |  |  |  |  |  |  |  |
| RHD associated with PUV | 19 | (3.3%) | 2 | (1.4%) | 76 | (5.1%) | 3 | (5.5%) |
| Solitary kidney | 24 | (4.2%) | 14 | (9.9%) | 204 | (13.8%) | 2 | (3.7%) |
| Bilateral RHD | 125 | (21.9%) | 38 | (26.8%) | 86 | (5.8%) | 12 | (22.2%) |
| Unilateral RHD | 111 | (19.5%) | 25 | (17.6%) | 147 | (9.9%) | 19 | (35.1%) |
| MCDK | 15 | (2.6%) | 11 | (7.7%) | 25 | (1.7%) | 12 | (22.2%) |
| Horseshoe kidney | 10 | (1.8%) | 0 | (0%) | 72 | (4.9%) | 0 | (0%) |
| Others<sup>b</sup> | 266 | (46.7%) | 52 | (36.6%) | 873 | (58.9%) | 6 | (11.1%) |
| Extrarenal anomalies, n (%) |  |  |  |  |  |  |  |  |
| Behavioral cognitive abnormality | 23 | (4.0%) | 11 | (7.7%) | 91 | (6.1%) | 4 | (7.4%) |
| Motor retardation | 10 | (1.8%) | 9 | (6.3%) | 152 | (10.2%) | 1 | (1.8%) |
| CHD | 30 | (5.3%) | 9 | (6.3%) | 174 | (11.7%) | 6 | (11.1%) |
| Skeletal malformation | 23 | (4.0%) | 10 | (7%) | 38 | (2.6%) | 2 | (3.7%) |
| Genitourinary malformation | 19 | (3.3%) | 5 | (3.5%) | 83 | (5.6%) | 1 | (1.8%) |
| CNS abnormality | 14 | (2.5%) | 9 | (6.3%) | 33 | (2.2%) | 1 | (1.8%) |
| Facial dysmorphism | 3 | (0.5%) | 5 | (3.5%) | 3 | (0.2%) | 0 | (0%) |
| Hearing abnormality | 13 | (2.3%) | 4 | (2.8%) | 13 | (0.9%) | 0 | (0%) |
| Ocular abnormality | 22 | (3.9%) | 6 | (4.2%) | 65 | (4.4%) | 7 | (12.9%) |

|  |  |  |  |  |
| --- | --- | --- | --- | --- |
| External ear malformation | 9 (1.6%) | 6 (4.2%) | 15 (1.0%) | 0 (0%) |
| Gastrointestinal tract malformation | 7 (1.2%) | 3 (2.1%) | 74 (5.0%) | 1 (1.8%) |
| CKD stage, n (%) |  |  |  |  |
| I | 395 (69.3%) | 80 (56.3%) | 782 (52.7%) | 27 (50.0%) |
| II | 50 (8.8%) | 22 (15.5%) | 260 (17.5%) | 8 (14.8%) |
| III | 52 (9.1%) | 14 (9.9%) | 203 (13.7%) | 14 (25.9%) |
| IV | 40 (7.0%) | 7 (4.9%) | 125 (8.4%) | 5 (9.2%) |
| V | 33 (5.8%) | 19 (13.4%) | 113 (7.6%) | 0 (0%) |
| Disease course, median (IQR) | 5.76 (3.88, 7.50) | 1.18 (0.25, 3.14) | 0.52 (0.03, 2.28) | 3.10 (1.76, 7.88) |

<sup>a</sup>Others include: *Robo2*, *LMX1B*, *NFIA*, *NSD1*, *SEC61A1* and *SETD5*. <sup>b</sup>Others include: ureteropelvic junction obstruction, ureterovesical junction obstruction, vesicoureteral reflux, duplex collecting system, hydronephrosis, and renal ectopia. CNV, copy number variation; CAKUT, congenital anomalies of the kidney and urinary tract; RHD, renal hypodysplasia; MCDK, multicystic dysplastic kidney; PUV, posterior urethral valves; CKD, chronic kidney disease; CHD, congenital heart disease; CNS, central nervous system; IQR, interquartile range.

### Model construction

In the derivation cohort, we developed a POCC model for predicting kidney failure at three time points (1, 3, and 5 years). A total of 30 candidate variables were collected (Supplementary Table 1), including baseline clinical features (n=8), renal phenotype (n=1), extrarenal phenotypes (n=11), and genetic features (n=10). To identify and retain only the most relevant features, we employed the XGBoost algorithm combined with SHAP (SHapley Additive exPlanations) analysis, computing the mean absolute SHAP values for all variables at the three prediction time points (1, 3, and 5 years), and ranked them (Supplementary Figure 1). We retained only features that did not have all-zero SHAP values at the three time points. Finally, we selected 9 key clinical phenotypic features (gender, age at first diagnosis, preterm birth, CKD stage at baseline, CAKUT subphenotype, short stature, congenital heart disease, ocular abnormality, and behavioral cognitive abnormality) to construct the POCC-general model. Given that CAKUT is a common congenital birth defect with a genetic etiology in 12–20% of cases, we incorporated three additional congenital hereditary features (CAKUT family history, prenatal phenotype, and *PAX2* mutation) to retain the 9 features above and further developed a POCC-specialized model containing 12 features to better tailor it to pediatric characteristics (Figure 2).

**Fig. 2:**
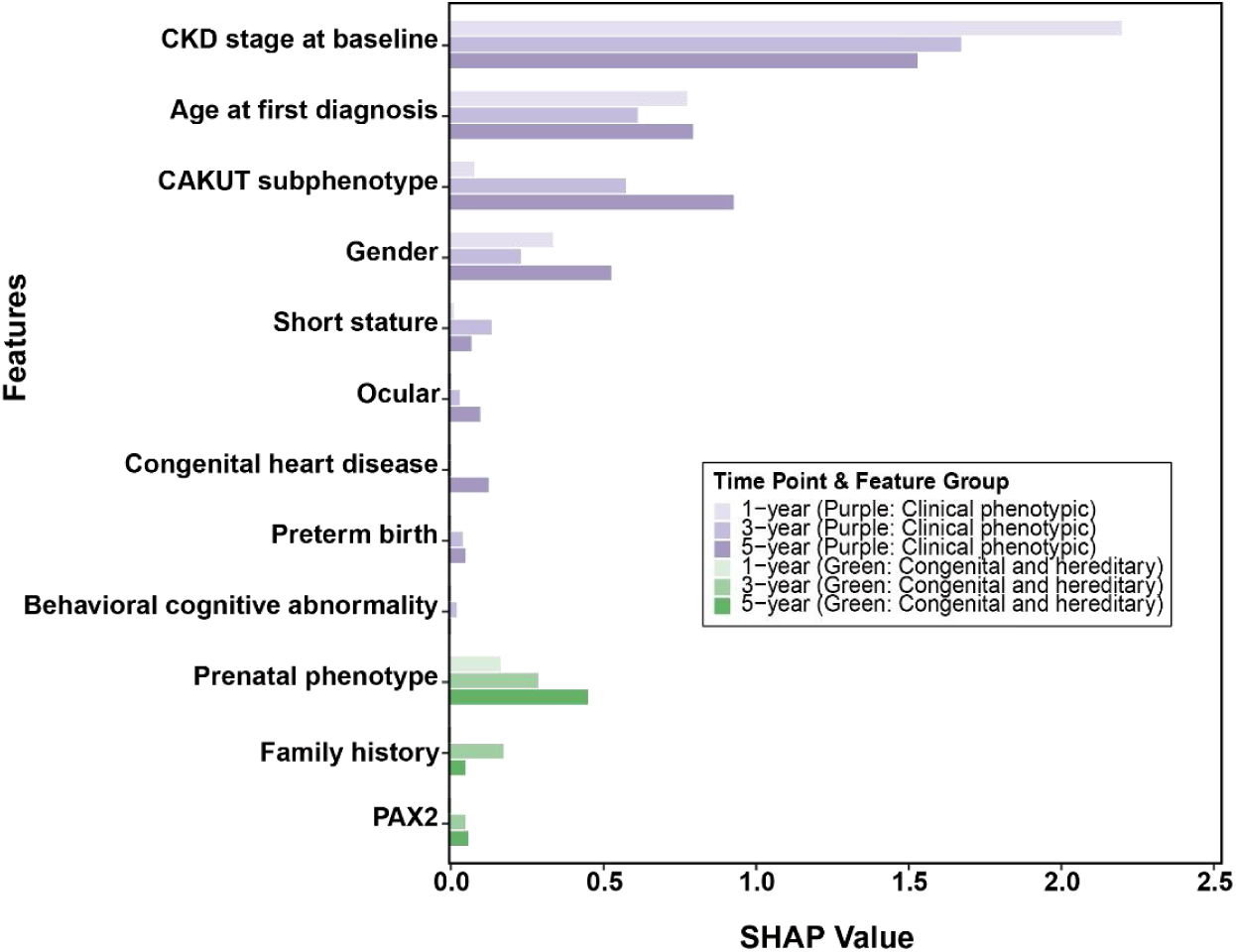
Top 12 features based on SHAP and XGBoost algorithms for kidney failure prediction at 1-, 3-, and 5-years. Purple coloring represents the 9 clinical phenotypic features, ranked in descending order by the cumulative sum of SHAP values across all three time points (1-year, 3-year, and 5-year predictions). Green coloring represents 3 congenital and hereditary features, arranged according to the same ranking criterion. The general model was constructed using only the 9 clinical-phenotypic features, whereas the specialized model incorporated all 12 features, combining the 9 clinical-phenotypic features with the 3 congenital and hereditary features. CAKUT, congenital anomalies of the kidney and urinary tract; CKD, chronic kidney disease; SHAP, SHapley Additive exPlanations. Others comprised the remaining 19 features with SHAP values of zero.

### Model comparison and external validation

To construct the POCC-general model, we utilized 9 selected key clinical phenotypic variables and developed 8 different ML algorithms, including eXtreme Gradient Boosting (XGBoost), Random Forest (RF), Support Vector Machine (SVM), K-Nearest Neighbors (KNN), Artificial Neural Network (ANN), Categorical Boosting (CatBoost), Gradient Boosting Machine (GBM), and Adaptive Boosting (AdaBoost). A comprehensive comparison of the performance of various algorithms (see Figure 3 and Supplementary Table 2) indicated that CatBoost achieved the best predictive performance. In the internal test cohort, CatBoost demonstrated excellent discriminative ability, with AUROCs at the 1-, 3-, and 5-year prediction time points of 0.988 (0.967–1.000), 0.972 (0.940–0.997), and 0.925 (0.836–0.982), respectively, and corresponding AUPRC values of 0.914 (0.725–1.000), 0.871 (0.719–0.978), and 0.831 (0.668–0.939), respectively. Moreover, the Brier scores at the three prediction time points ranged from 0.027 to 0.120, indicating good calibration. To further validate the generalizability of CatBoost, two multicenter independent external validation cohorts were used. In external validation cohort 1, comprising 14 pediatric specialty hospitals from the CCGKDD, CatBoost demonstrated AUROC values ranging from 0.895 to 0.980, AUPRC values from 0.914 to 0.954, and Brier scores from 0.071 to 0.214 across the three prediction time points. In external validation cohort 2, which comprised 26 general hospitals from the CRDS, the AUROC values at all three prediction time points were ≥0.80 (with the 1-year AUROC reaching 0.897), the AUPRC values ranged from 0.789 to 0.884, and the Brier scores ranged from 0.088 to 0.212, demonstrating robustness. Beyond the aforementioned metrics, CatBoost demonstrated strong performance across accuracy, precision, sensitivity, specificity, and F1 score (see Supplementary Figure 2). Decision curve analysis further validated that the model provided satisfactory clinical net benefit (Supplementary Figure 3).

**Fig. 3:**
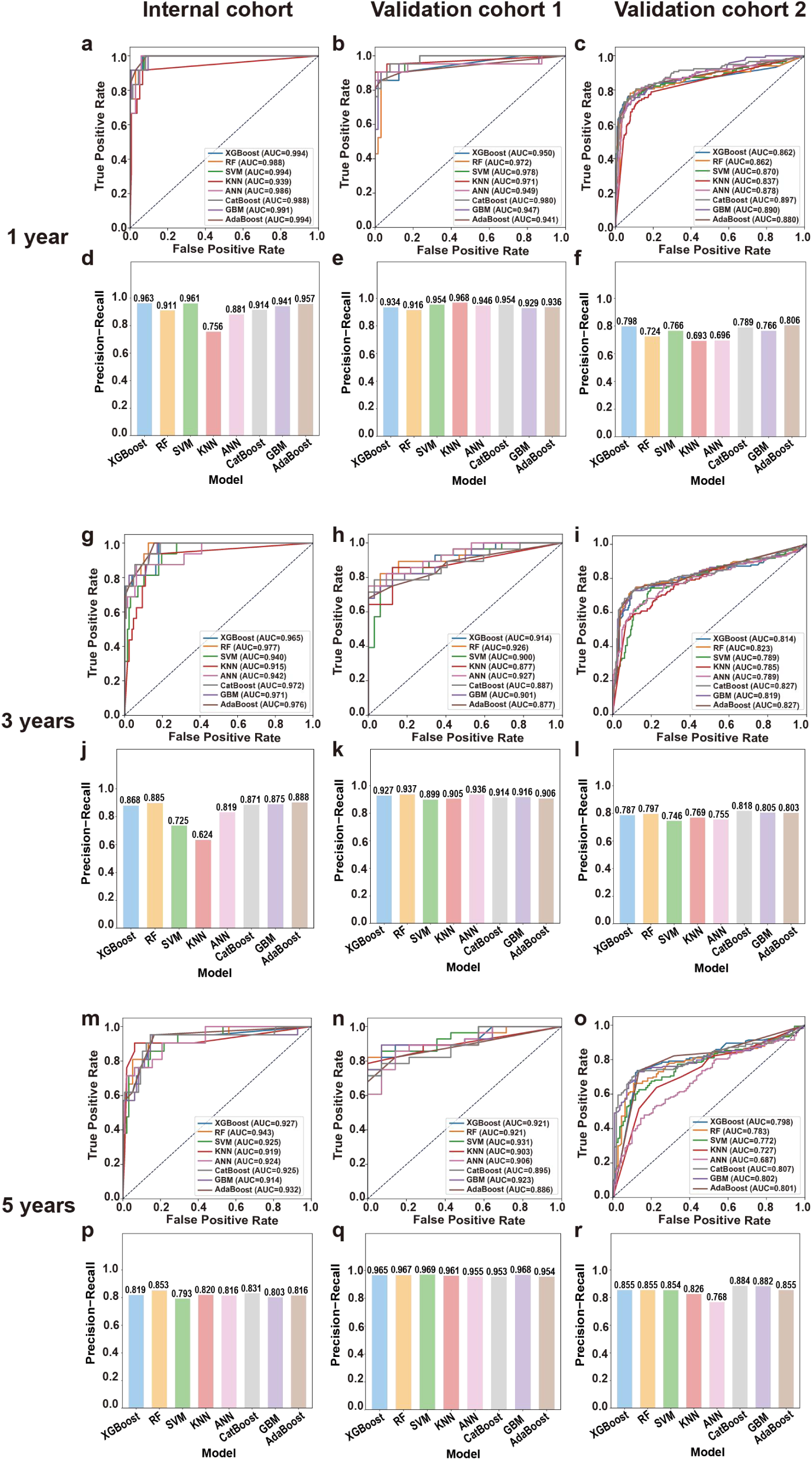
Performance of the general model across eight machine learning algorithms in the internal test and external validation cohorts. Panels a–c, g–i, and m–o display AUROC curves for 1-, 3-, and 5-year risk prediction, respectively. Panels d–f, j–l, and p–r show the corresponding AUPRC for the same prediction time points. XGBoost, extreme gradient boosting; RF, random forest; SVM, support vector machine; KNN, k-nearest neighbors; ANN, artificial neural network; CatBoost, categorical boosting; GBM, gradient boosting machine; AdaBoost, adaptive boosting.

In constructing the POCC-specialized model, the variables included the aforementioned 9 clinical phenotypic variables combined with 3 congenital and hereditary variables, and the same 8 ML algorithms were applied. Comparisons of various algorithms showed that the CatBoost algorithm consistently achieved the best predictive performance in this task (Figure 4 and Supplementary Table 3). In the internal test cohort, the specialized model’s AUROC ranged from 0.931 to 0.993, its AUPRC ranged from 0.833 to 0.950, and its Brier scores ranged from 0.034 to 0.111. Because complete congenital and hereditary variables were only available in the CCGKDD, external validation of the specialized model was performed solely in external validation cohort 1. The results demonstrated that CatBoost had excellent performance across all metrics in this external validation cohort. The AUROC values at the three prediction time points ranged from 0.908 to 0.960, the AUPRC values ranged from 0.929 to 0.959, and the Brier scores ranged from 0.059 to 0.143. Compared with the general model’s performance in this cohort, the specialized model demonstrated superior calibration and comparable discriminative ability. Additionally, the specialized model exhibited improved accuracy (0.833–0.941), precision (0.875–0.958), sensitivity (0.750–0.857), specificity (0.906–0.969), and F1 score (0.808–0.885) (see Figure 4). Decision curve analysis was used to quantify the clinical value of the specialized model. In both the internal test cohort and the external validation cohort 1, when the threshold probability ranged from 0.10–0.65 and 0.28–0.80, respectively, CatBoost demonstrated satisfactory net clinical benefit across the three time points compared with the “treating all” or “treating none” strategies (Supplementary Figure 4).

**Fig. 4:**
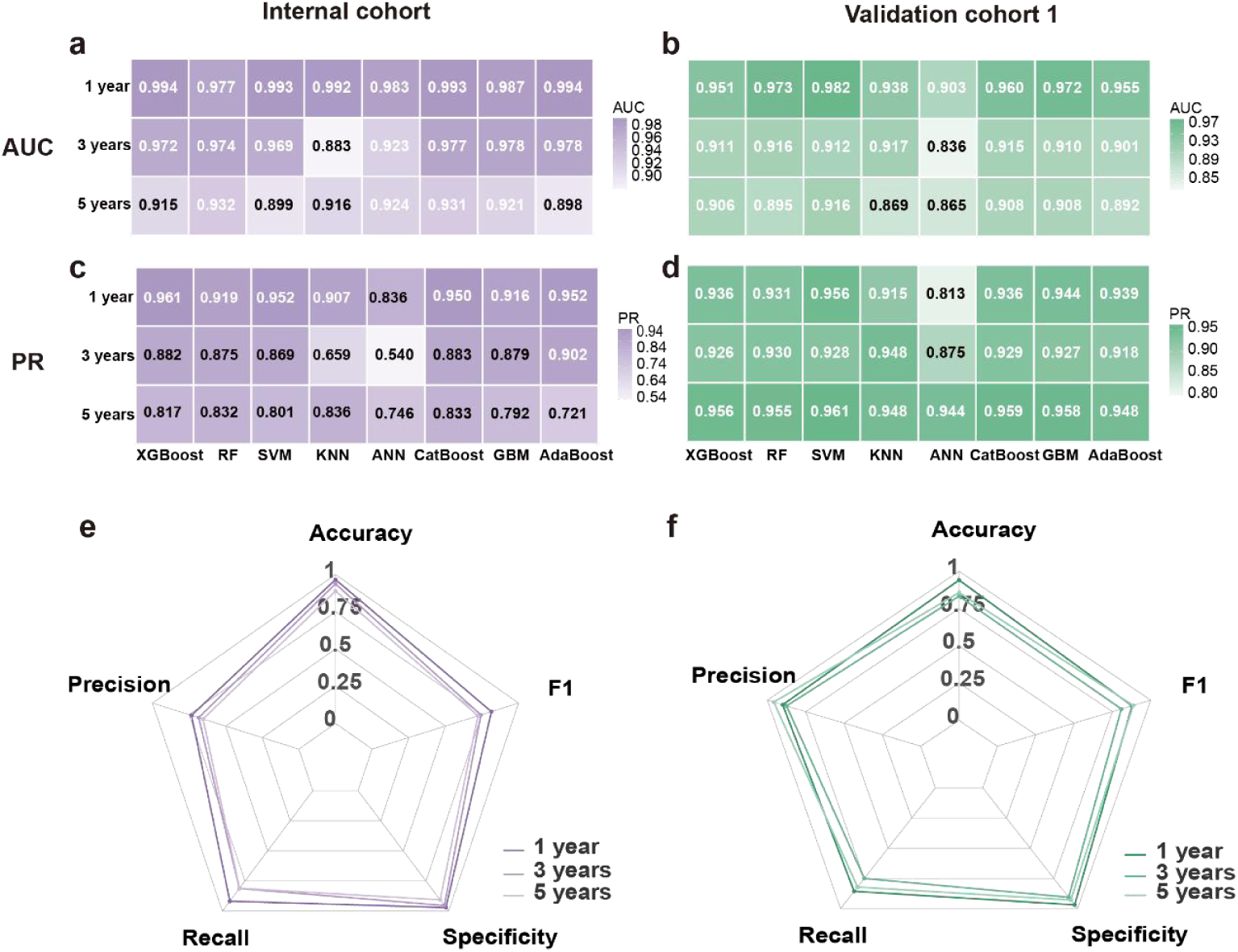
Performance of the specialized model in the internal test and external validation cohorts. Panels a–b show AUROC heatmaps and panels c–d show AUPRC heatmaps for internal test and external validation cohorts, respectively. Panels e–f present radar plots summarizing additional performance metrics (accuracy, precision, recall, specificity, and F1-score) for internal test and external validation cohorts, respectively. XGBoost, extreme gradient boosting; RF, random forest; SVM, support vector machine; KNN, k-nearest neighbors; ANN, artificial neural network; CatBoost, categorical boosting; GBM, gradient boosting machine; AdaBoost, adaptive boosting.

### Optimal model interpretation

To enhance model interpretability, we employed SHAP analysis to quantify each variable’s contribution to CatBoost predictions. Figure 5 presents the SHAP summary plot for the general model, which visualizes the SHAP values for the nine key clinical phenotypic variables ranked by descending importance. Positive SHAP values indicate increased kidney failure risk. The summary plot revealed that CKD stage at baseline, CAKUT subphenotype, and age at first diagnosis consistently ranked among the top predictors at 1-, 3-, and 5-year time points, all of which exerting similar directional effects on kidney failure risk. We also constructed SHAP dependence plots to illustrate how individual features influence predictions. Specifically, kidney failure risk remained stable when the CKD stage at baseline was <3, but increased significantly at stage ≥ 3. Among CAKUT subphenotypes, RHD associated with PUV, bilateral RHD, and solitary kidney demonstrated significantly elevated positive SHAP values. Age at first diagnosis exhibited a nonlinear relationship with kidney failure risk: no significant increase during preschool years, but progressive elevation during school age and beyond (≥ 6 years). Additionally, we generated Sankey plots to compare feature importance rankings between general and specialized models. The results demonstrated that CKD stage at baseline, CAKUT subphenotype, and age at first diagnosis consistently ranked among the top three predictors in the specialized model across all time points. This high concordance with the general model further validates the robustness of these three features (Figure 5).

**Fig. 5:**
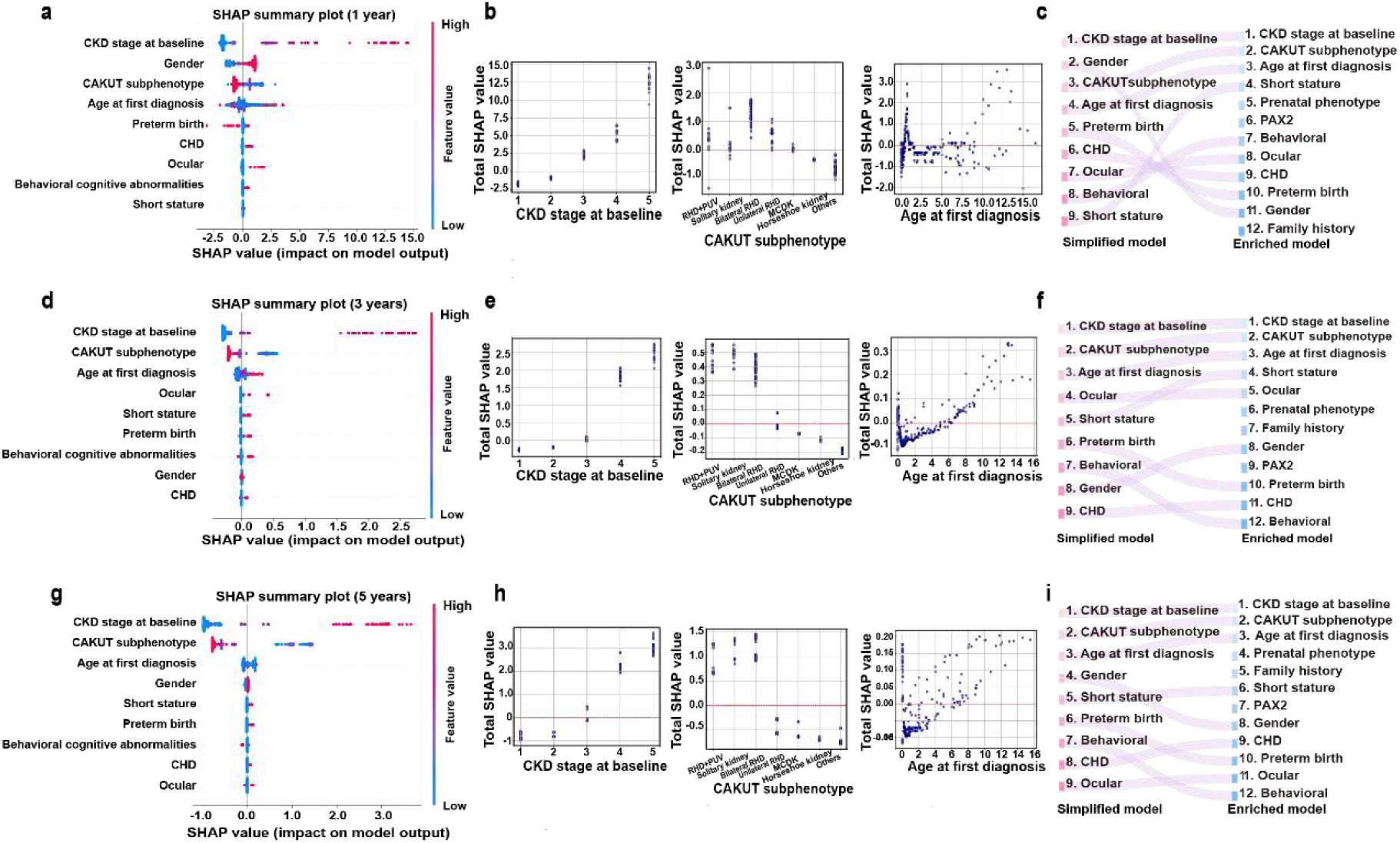
Model interpretability using SHAP. Panels a, d, and g present SHAP summary plots illustrating feature importance for 1-, 3-, and 5-year risk prediction in the general model, respectively. The corresponding SHAP dependence plots for the three time points are shown in panels b (1-year), e (3-year), and h (5-year), focusing on the top three influential features: CKD stage at baseline, CAKUT subphenotype, and age at first diagnosis. Sankey diagrams in panels c (1-year), f (3-year), and i (5-year) depict the temporal shifts in feature importance rankings between the general and specialized models across different prediction horizons. In the summary plot, each row corresponds to a feature, and each dot represents a single patient’s SHAP value. The x-axis displays the SHAP value, reflecting the magnitude and direction of the feature’s contribution to kidney failure (positive values indicate increased kidney failure risk, while negative values indicate decreased risk). The color gradient of the dots denotes the original feature value, ranging from low (blue) to high (red). Features are ranked from top to bottom along the y-axis by their total SHAP values. A wider spread along the x-axis indicates a greater overall impact of that feature on the model prediction. SHAP, SHapley Additive exPlanations.

### Model deployment and real-world evaluation

To facilitate clinical application and translation, we deployed POCC as a Streamlit-based interactive web application (https://kidneyfailurepredictionsystem.streamlit.app/). Clinicians can flexibly select either the specialized or the general model based on feature availability. After the relevant patient feature values are input, the application automatically estimates the individual risk of kidney failure at 1, 3, and 5 years after the initial CAKUT diagnosis. Additionally, the application generates SHAP force plots for each prediction time point to visualize individual feature contributions: blue features drive the prediction probability toward “no renal failure,” whereas red features drive it toward “renal failure” (Figure 6a).

**Fig. 6:**
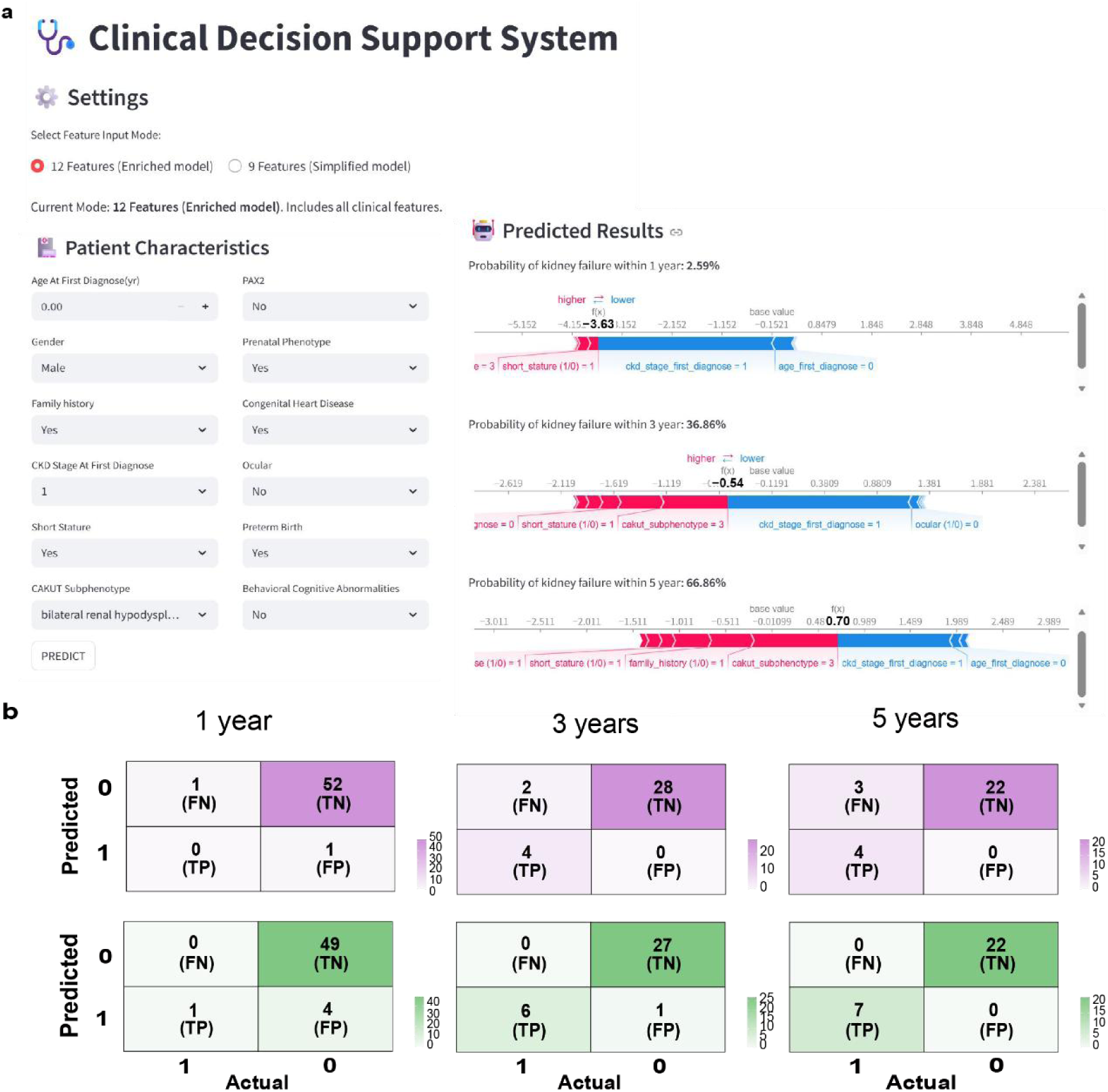
Online prediction platform and real-world validation performance. Panel a displays the user interface of the web-based prediction tool, enabling clinicians to flexibly select between the specialized model (12 features) and the general model (9 features) based on clinical context and data availability. Meanwhile, the force plot for an individual child indicates the features that contribute to the decision of “kidney failure”: the blue features on the right are the features pushing the prediction towards the “non-kidney failure” class, while the red features on the left are pushing the prediction towards the “kidney failure” class. Panel b shows confusion matrices comparing the performance of the general model (purple) and specialized model (green) for 1-, 3-, and 5-year kidney failure risk prediction in real-world validation. FN, false negative; FP, false positive; TN, true negative; TP, true positive.

Furthermore, we retrospectively collected data from 54 CAKUT patients from the Children’s Hospital of Fudan University (CHFU) registered in the CCGKDD and performed real-world validation using the web-based application to compare the performance of the general and specialized models (Figure 6b). For the general model, the 1-year prediction accuracy was 96.3% (1 false negative and 1 false positive), the 3-year prediction accuracy was 94.1% (2 false negatives), and the 5-year prediction accuracy was 89.7% (3 false negatives). Across all three prediction timepoints, the general model had 5 prediction errors (1 false positive and 4 false negatives), with an overall accuracy of 90.7%. In contrast, the specialized model achieved accuracies of 92.6% (4 false positives), 97.1% (1 false positive), and 100% (sensitivity and specificity both 100%) for 1-, 3-, and 5-year predictions, respectively. Across all three time points, the model incorrectly predicted 5 patients, yielding the same overall accuracy of 90.7%; however, all errors were false positives, and there were no false negatives.

### Sensitivity analysis

To validate the robustness of the model, we performed sensitivity analyses on the general model by collapsing three specific extrarenal phenotype variables (congenital heart disease, ocular abnormality, and behavioral cognitive abnormality) into a single binary extrarenal phenotype variable. Results demonstrated that the CatBoost-based general model maintained stable performance across the internal test and two external validation cohorts, with AUC fluctuations within 0.02 (Supplementary Figure 5). For the specialized model, we replaced the *PAX2* mutation variable with an aggregated binary genetic variable based on the aforementioned aggregation of extrarenal phenotype variables. Model performance remained essentially unchanged, with AUC fluctuations within 0.01 for both internal testing and external validation cohorts (Supplementary Figure 6), indicating excellent robustness and generalizability for both models.

## Discussion

In our multicenter study, we present POCC, an advanced ML-based model developed to establish a risk stratification tool for kidney failure at 1, 3, and 5 years after initial diagnosis, using baseline clinical information, renal and extrarenal phenotypes, and genetic data. Our study is the first to incorporate multiple subphenotypes and time points into CAKUT kidney failure prediction and is also the largest in scale among similar studies. POCC demonstrated robust performance in the derivation and two independent external validation cohorts (AUC ≥ 0.90) and maintained 90.7% accuracy in real-world validation. The development of general and specialized models further enhanced clinical generalizability and applicability. Furthermore, SHAP analysis revealed the three most important predictors and elucidated their nonlinear associations with outcomes, which provides novel insights for individualized intervention. Our study indicates that POCC may serve as a potential predictive tool for assessing kidney failure progression in children with CAKUT and may help clinicians achieve precise identification of patients at high risk, facilitate timely individualized interventions to slow kidney function decline, and support preparation for renal replacement therapy.

In recent years, advances in algorithms have enabled the widespread adoption of real-world data-driven clinical modeling for classification, prediction, and evidence-based decision-making^16^. Douglas et al.^11^ developed a binary logistic regression model to predict kidney failure within 6 months of CAKUT diagnosis. Similarly, Isabel et al.^12^ applied a Cox proportional hazards model to assess the risk of renal function decline in patients with CAKUT. However, traditional statistical models typically rely on population-level characteristics and incorporate only a limited number of variables, thereby falling short of modern precision medicine’s requirements for individualized risk prediction. To our knowledge, only two prior studies have developed ML models for predicting kidney failure in specific CAKUT subphenotypes. Jethro et al.^14^constructed a random survival forest model for predicting CKD progression, initiation of kidney replacement therapy, and clean intermittent catheterization in patients with posterior urethral valves. Their external validation cohort achieved a C-index of 0.89, thus outperforming conventional Cox regression. However, the small sample size (103 training, 22 testing) posed a risk of overfitting, and the composite outcome included subjective catheterization thresholds, potentially limiting generalizability. Richter et al.^15^developed a random forest model to predict the risk of postnatal kidney failure or death in fetuses with prenatally diagnosed lower urinary tract obstruction and achieved 0.77 accuracy. Nevertheless, the single-center design and lack of external validation limit its applicability. Beyond kidney failure endpoints, existing studies have primarily addressed non-kidney failure outcomes, such as surgical intervention or spontaneous resolution, and have been restricted to specific CAKUT subphenotypes, including vesicoureteral reflux, prenatal hydronephrosis, and pelviureteric junction obstruction^17–19^. Critically, to date, no prediction model has been developed for multiple CAKUT subphenotypes. In this study, we addressed this gap by recruiting the largest CAKUT dataset to date (n=2,249) from 41 centers in the CCGKDD and CRDS, encompassing twelve subphenotypes, including posterior urethral valves, renal dysplasia, solitary kidney, and horseshoe kidney. To assess generalizability, reliability, and effectiveness, we additionally recruited 142 CAKUT children from 14 pediatric specialty hospitals across 9 provinces in China as external validation cohort 1 and 1,483 children from 26 general hospitals across seven regions, including Northwest, Central, South, Southwest, East, Northeast, and North China, as external validation cohort 2. We validated the POCC model across two distinct hospital dimensions—the pediatric specialty hospital cohort and general hospitals—and achieved external AUCs of 0.98 and 0.90, respectively, and demonstrated robust temporal generalization across 1-, 3-, and 5-year time windows. More importantly, to support clinical implementation, we developed a user-friendly online platform designed for convenient clinical use. This platform enables clinicians to obtain immediate kidney failure risk predictions upon inputting predictive factors. In a retrospective real-world cohort of 54 patients from the CHFU, the platform achieved an impressive 90.7% accuracy. These findings demonstrate that POCC has excellent cross-institutional reproducibility, temporal stability, and geographic generalizability, with strong potential to support translation in real-world clinical settings.

The flexible model development strategy represents another advantage of this study. We developed and validated both general and specialized models to accommodate implementation needs across different healthcare settings. The general model, which was constructed using baseline clinical information and renal and extrarenal phenotypic variables—all routinely available clinical metrics—offers particular value for deployment in primary care institutions or general hospitals. Inclusion of extrarenal phenotypic features, such as congenital heart disease and ocular abnormalities, enables coverage and benefit for patients with CAKUT syndromes. Furthermore, the general model achieved AUCs of 0.980 and 0.897 in two independent external validation cohorts from pediatric specialty hospitals and general hospitals, respectively, while maintaining 90.7% prediction accuracy in the real-world validation cohort. These results demonstrate that the general model offers high cost-effectiveness and broad applicability, providing rapid decision-making support in diverse healthcare settings. On the other hand, considering the nature of CAKUT as a congenital and hereditary disease, we developed a specialized model more tailored to CAKUT and pediatric characteristics by adding three features: family history of CAKUT, prenatal phenotype, and genetic feature (*PAX2* mutation). The specialized model achieved an AUC of 0.960 in the externally validated pediatric specialty hospital cohort and similarly performed well, with 90.7% accuracy in real-world validation. Notably, the two models exhibited different error patterns in real-world settings: the specialized model produced exclusively false-positive predictions (risk overestimation), whereas the general model generated primarily false-negative predictions (risk underestimation). In life-threatening conditions such as kidney failure, the risk of treatment delay from false-negative predictions may far outweigh the harm of excessive anxiety from false-positive predictions^20^. Therefore, the specialized model may provide greater clinical safety. Moreover, sensitivity analysis indicated that in real clinical scenarios, when patients present with extrarenal phenotypes beyond those explicitly included or with gene mutations other than *PAX2*, clinicians need only update the binary variables indicating whether extrarenal phenotypes or gene mutations are present, and both models maintain robust predictive performance. These findings validate their flexibility across diverse clinical phenotypes and genetic backgrounds.

Furthermore, interpretability and transparency of POCC are essential for gaining clinicians’ trust^21^. We employed SHAP to interpret the model, rank feature importance, and elucidate nonlinear relationships between features and outcomes. The results demonstrated that CKD stage at baseline ≥3, specific CAKUT subphenotypes (RHD associated with PUV, bilateral renal dysplasia, and solitary kidney), and age at first diagnosis ≥6 years contributed most to kidney failure prediction. These key features were validated across the horizontal dimension of general versus specialized models and the longitudinal dimension of 1-, 3-, and 5-year timepoints. CKD stage at baseline ≥ 3, and the three specific CAKUT subphenotypes have been previously established as risk factors^22–25^. Age at first diagnosis ≥6 years represents a novel finding of this study, which may reflect the higher prevalence of advanced CKD among older children at presentation and the accelerated loss of kidney function associated with pubertal development^22^. A large North American Pediatric Renal Trials and Collaborative Studies cohort previously identified age ≥ 5 years at diagnosis as a critical threshold associated with a 1.3- to 1.9-fold increased risk of kidney failure^26^, corroborating our findings. These results suggest that shifting the clinical intervention window to before school age could improve outcomes for CAKUT patients and indicate the importance of early postnatal urinary system ultrasound screening^27^.

However, this study has several limitations. First, the model was constructed from a long-term retrospective cohort, which may have introduced variation in information collection, thereby limiting its applicability. Second, because the data were sourced from multiple centers across China, certain variables (such as proteinuria and hypertension) were difficult to capture, and serial longitudinal clinical data across various follow-up assessments were not systematically recorded. Third, although integrating the SHAP algorithm with the ML model enhanced interpretability, increased clinician trust, and facilitated the extraction of meaningful clinical insights, clinical judgment cannot be entirely replaced. In the future, incorporating multimodal data, including imaging data, may further support the development and validation of more robust predictive models.

In conclusion, we developed POCC using ML algorithms to precisely predict kidney failure risk per patient at 1, 3, and 5 years after initial CAKUT diagnosis. The POCC demonstrated robust predictive performance in two independent external validation cohorts spanning both pediatric specialty and general hospitals and showed excellent temporal generalizability across all three prediction timepoints. Notably, the POCC-specialized model achieved better discrimination and reliability in real-world settings based on the online platform, demonstrating strong potential for translation. In clinical practice, POCC holds promise as a practical tool for precise, patient-specific assessment of kidney failure risk over time in CAKUT children, facilitating the implementation of tailored follow-up management plans and timely interventions and ultimately improving patient outcomes.

## Methods

### Study population

This multicenter retrospective study included four different cohorts derived from two large national clinical databases. Study samples were sourced from the Chinese Children Genetic Kidney Disease Database (CCGKDD) and the China Renal Data System (CRDS). The CCGKDD, which is led by the National Children’s Medical Center of China (Children’s Hospital of Fudan University), was established in 2014 and officially launched in September 2017. It is a genotype-phenotype database for pediatric hereditary kidney diseases that integrates clinical data from 146 pediatric specialty hospitals and pediatric nephrology departments across general hospitals nationwide^28^. The CRDS was jointly established by the National Clinical Research Center for Kidney Disease and the Chronic Disease Center of the Chinese Center for Disease Control and Prevention in 2018 and integrates digital electronic health records from 10.29 million hospitalized patients across 28 hospitals nationwide (including 1 pediatric specialty hospital and 27 general hospitals)^29,30^.

A total of 2,249 patients admitted between January 2014 and October 2025 were enrolled in this study, and were stratified into four cohorts: (1) derivation cohort (n=570, from the CHFU in the CCGKDD); (2) external validation cohort 1 (n=142, from 14 other pediatric specialty hospitals excluding CHFU in the CCGKDD); (3) external validation cohort 2 (n=1,483, from 26 general hospitals in the CRDS); and (4) real-world cohort (n=54, from the CHFU in the CCGKDD). Inclusion criteria were as follows: (1) age ≤18 years and (2) a diagnosis of CAKUT confirmed by ultrasound or other imaging modalities. Exclusion criteria included patients whose baseline serum creatinine, height, or follow-up outcome information was missing. This study strictly adhered to the Transparent Reporting of a multivariable prediction model for Individual Prognosis or Diagnosis (TRIPOD) guidelines and was approved by the Ethics Committee of the CHFU (No. 2022-05). Patients from the CCGKDD provided written informed consent and the informed consent was waived for patients from CRDS. Detailed information on the study population is shown in Figure 7.

**Fig. 7:**
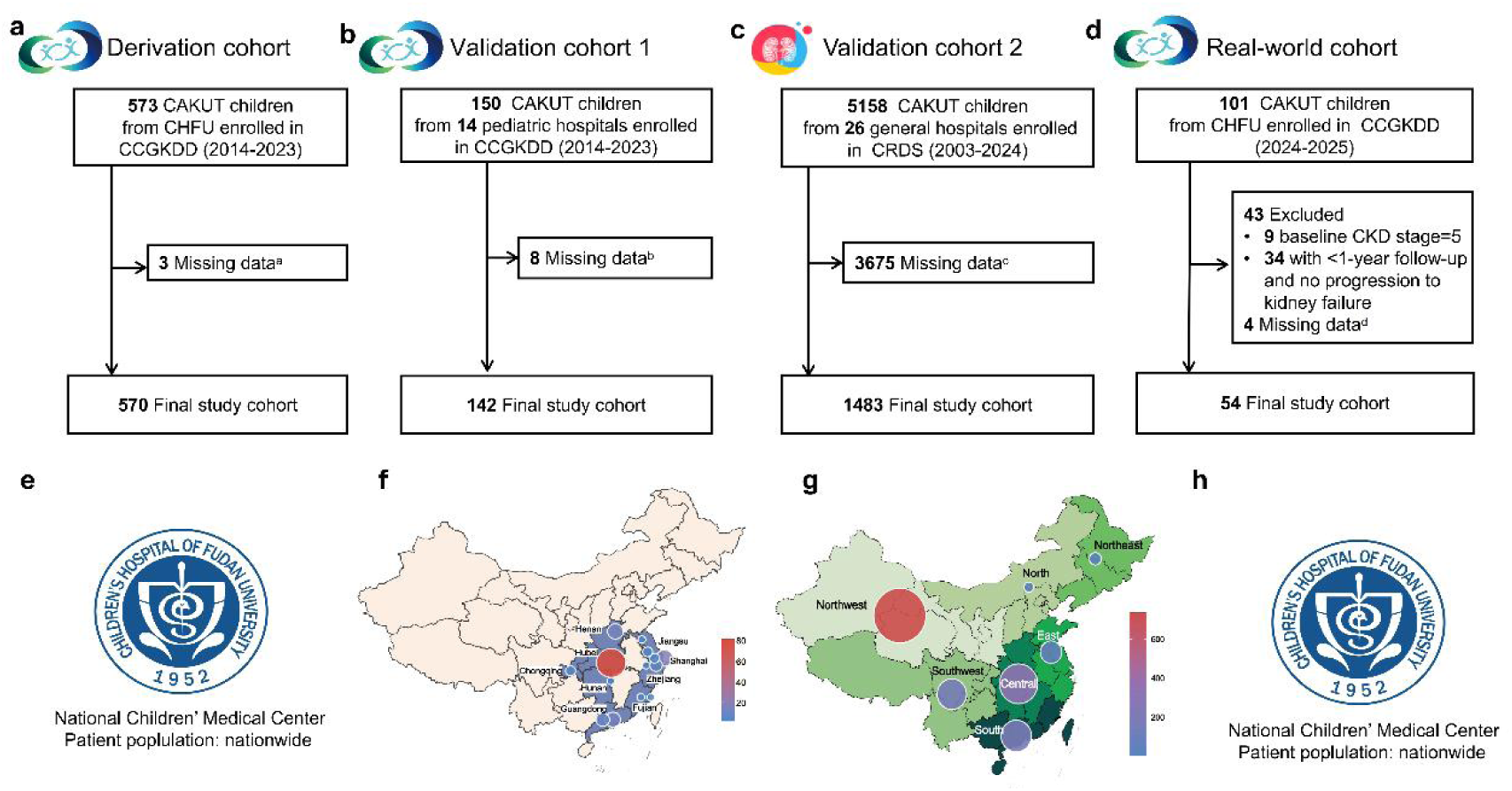
Study flow. Panel a illustrates the derivation cohort, recruited from CHFU (patients were recruited from 27 provinces, municipalities, and autonomous regions across China) within the CCGKDD (panel e). Panel b represents external validation cohort 1, comprising patients from 14 pediatric specialty hospitals across nine provinces (including Hubei, Guangdong, Shanghai, Henan, Zhejiang, Jiangsu, Chongqing, Fujian, and Hunan) in the CCGKDD (panel f). Panel c shows external validation cohort 2, sourced from 26 general hospitals across seven regions (including Northwest, Central, South, Southwest, East, Northeast, and North China) within the CRDS (panel g). Panel d depicts the real-world validation cohort, derived from CHFU within the CCGKDD (panel h). ^a^3 cases without baseline serum creatinine; ^b^8 cases without baseline serum creatinine; ^c^1,670 cases without baseline serum creatinine and 2,005 cases without height data; ^d^4 cases with outcome date not documented. CAKUT, congenital anomalies of the kidney and urinary tract. CHFU, Children’s Hospital of Fudan University; CCGKDD, Chinese Children Genetic Kidney Disease Database; CRDS, China Renal Data System; CKD, chronic kidney disease.

### Data collection and preprocessing

The candidate variables in this study included baseline clinical information, renal phenotypes, extrarenal phenotypes, and genetic information, which were systematically collected based on the relevant literature on risk factors and consultations with pediatric nephrology specialists. All data were retrospectively collected from electronic medical records. Baseline clinical features included gender, age at first diagnosis, preterm birth, prenatal phenotype, family history of CAKUT, CKD stage at baseline, short stature, and hyperuricemia. Renal phenotypes were classified according to the scheme proposed by Simone et al.^31^, which covers 12 CAKUT subphenotypes: renal hypodysplasia (RHD) associated with posterior urethral valves (PUV), solitary kidney, bilateral RHD, unilateral renal RHD, multicystic dysplastic kidney (MCDK), horseshoe kidney, and other types (including ureteropelvic junction obstruction, ureterovesical junction obstruction, vesicoureteral reflux, duplex collecting system, hydronephrosis, and renal ectopia). When multiple CAKUT subphenotypes coexisted, the patient was classified according to the subphenotype with the most significant effect on renal function degradation. For instance, patients with RHD and VUR were stratified as RHD. Extrarenal phenotypes included congenital heart disease, ocular abnormality, behavioral cognitive abnormality, motor retardation, skeletal malformation, genitourinary malformation, central nervous system abnormality, facial dysmorphism, hearing abnormality, external ear malformation, and gastrointestinal malformation. Genetic testing methods in the CCGKDD included targeted exome sequencing, proband whole-exome sequencing, and trio whole-exome sequencing. The genetic variables included copy number variants (CNVs), chromosomal abnormalities, and monogenic mutations (e.g., *PAX2*, *TNXB*, *EYA1*, *HNF1β*, *GATA3*, *SALL1*, *COL4A1*). With the exception of CKD stage at baseline, all the other variables were collected at the time of initial CAKUT diagnosis. CKD stage at baseline was calculated based on the most recent serum creatinine level within 6 months before or after the CAKUT diagnosis date. The estimated glomerular filtration rate (eGFR) was calculated using the updated Schwartz equation^32^. A family history of CAKUT was defined as the presence of CAKUT among blood relatives within three generations. Prenatal phenotype was defined as abnormal urinary tract structures or oligohydramnios detected on prenatal fetal ultrasound. The primary outcome was kidney failure occurrence within 1, 3, and 5 years after diagnosis, with kidney failure defined as eGFR <15 mL/min/1.73 m² or initiation of kidney replacement therapy. Detailed definitions of other variables are provided in Supplementary Table 1.

During data preprocessing, extreme outliers from the CCGKDD were flagged and referred to clinicians for reevaluation, and missing values were supplemented by clinicians through review of electronic medical records or telephone follow-up. For the CRDS, structurally extracted data had been previously cleaned, standardized, anonymized, and integrated. Missing data were handled as follows. Patients with missing baseline serum creatinine level, height, or outcome information were excluded. For other variables (such as preterm birth, short stature, and extrarenal phenotypes), if no relevant records were found in either ICD-10 codes or electronic medical records, then they were uniformly coded as normal. Additionally, patients with an incomplete follow-up (i.e., those whose actual follow-up time did not reach the target prediction horizon and who did not experience kidney failure during that period) were excluded to ensure the accuracy and completeness of the study data, as detailed in Supplementary Figure 7.

### Model development

Model development encompasses two core components: variable selection and algorithm optimization. For feature selection, XGBoost integrated with the SHAP algorithm was used to calculate the mean absolute SHAP values for all candidate variables across the 1-year, 3-year, and 5-year prediction horizons. Key features were selected based on the criterion that the variables did not demonstrate all-zero SHAP values across the three time points, ultimately establishing a robust variable combination applicable to all the prediction horizons (Supplementary Figure 1). To flexibly construct the Predicting Outcome in Children with CAKUT (POCC) model, the selected variables were stratified into clinical phenotypic as well as congenital and hereditary features. The former independently constructed the POCC-general model, while their combination constructed the POCC-specialized model. For algorithm optimization, eight ML algorithms were employed: eXtreme Gradient Boosting (XGBoost), Random Forest (RF), Support Vector Machine (SVM), K-Nearest Neighbors (KNN), Artificial Neural Network (ANN), Categorical Boosting (CatBoost), Gradient Boosting Machine (GBM), and Adaptive Boosting (AdaBoost). Hyperparameter tuning was implemented via a grid search with 10-fold cross-validation (Supplementary Tables 4 and 5), and early stopping was introduced to prevent overfitting. Dataset partitioning employed a stratified strategy: algorithms using early stopping were allocated training, validation, and test sets at a 6:1:3 ratio, while remaining algorithms were split into training and test sets at a 7:3 ratio.

### Model evaluation and explanation

Final prediction models were constructed using the optimal feature combinations and eight algorithms, with performance evaluated across three dimensions: discrimination, calibration, and clinical utility. Discrimination was assessed using the area under the receiver operating characteristic curve (AUROC) and area under the precision-recall curve (AUPRC), with the AUROC as the primary metric, supplemented by accuracy, precision, recall, specificity, and F1 score. Calibration was quantified using the Brier score^33^, while clinical utility was evaluated through decision curve analysis (DCA) to assess clinical net benefit across different decision thresholds^34^. Internal validation was performed using 1,000 bootstrap resamples of the training cohort. External validation was conducted across two independent multicenter cohorts from CCGKDD and CRDS for the general model and within the CCGKDD cohort exclusively for the specialized model. SHAP analysis was employed to interpret the underlying prediction mechanisms^35^. SHAP summary plots identified and ranked key predictors and illustrated the global importance of each feature. SHAP dependence plots elucidated the relationships between individual predictors and outcomes, whereas SHAP local explanation plots illustrated each feature’s specific contribution to individual predictions. Sankey diagrams were employed to compare the evolution of feature importance between the general and specialized models.

### Model deployment and real-world study

To enhance clinical applicability, we developed a user-friendly, interactive web application using the Streamlit framework. The application simultaneously hosts both the specialized and general models, allowing users to flexibly select them based on feature availability. It provides real-time predictions of kidney failure risk at 1, 3, and 5 years after initial diagnosis and visualizes each feature’s contribution to model decision-making through SHAP force plots. Based on this online prediction tool, 101 CAKUT patients admitted to CHFU and registered in the CCGKDD between January 2024 and October 2025 were retrospectively enrolled. After the same inclusion and exclusion criteria were applied, 54 patients were ultimately included in the real-world validation study (Fig. 7d). The performance in the real-world validation was evaluated using confusion matrices.

### Sensitivity analysis

To comprehensively evaluate model robustness, we conducted sensitivity analyses using the following variable merging and replacement strategies. (1) In the general model, three extrarenal phenotype variables (congenital heart disease, ocular abnormality, and behavioral cognitive abnormality) were merged into a binary variable indicating presence or absence of extrarenal phenotypes. (2) In the specialized model, in addition to merging the aforementioned three extrarenal phenotypes, *PAX2* mutation was replaced with a binary variable indicating the presence or absence of any genetic variant (Supplementary Figure 8).

### Statistical analysis

Normality was assessed for continuous variables. Normally distributed variables are presented as mean ± standard deviation, whereas nonnormally distributed variables are shown as median (interquartile range). Between-group comparisons of parametric continuous variables were performed using independent-samples Student’s t tests, whereas nonparametric variables were compared with the Mann–Whitney U test. Categorical data are reported as frequency (percentage) and compared via the chi-square test or Fisher’s exact test, as appropriate. All statistical tests were two-sided, with *P* < 0.05 considered to indicate statistical significance. A list of the packages used for the analyses (Python version 3.13.2 and R version 4.5.0) is provided in Supplementary Table 6.

## Data availability

All data generated or analysed during this study are included in this published article and its supplementary information files.

## Code availability

Code is available upon request from the corresponding authors.

## Consent to participate

This study was approved by the Ethics Committee of the CHFU (No. 2022-05). Patients from the CCGKDD provided written informed consent and the informed consent was waived for patients from CRDS.

## Clinical trial number

not applicable.

## Acknowledgement

Shanghai Municipal Education Commission Program on AI for Transforming Academic Research Paradigms (24RGZNB05); Fudan University Medicine-Engineering Collaboration Program (yg-2025-general-07); The Medicine-X Innovation Team Incubation Program at the Children’s Hospital of Fudan University (EKYX202413); The National Key Research and Development Program (2021YFC2500202); China Kidney Disease Big Data Collaboration Network Open Research Program (CRDS-004016).

## Author contributions

**T.L., H.W., J.L.** Data curation, Formal analysis, Software, Validation, Visualization, Writing–Original Draft

**X.Z., Y.X., X.W., Y.K., C.L., X.G., X.J., J.M., Y.L.(Yufeng Li), A.Z., M.W., H.B., T.S., X.D., D.W., R.Z., Y.L.(Yihan Lu), Q.S.** Data curation, Methodology.

**S.N.** Conceptualization, Data curation, Methodology, Supervision, Writing–Review & Editing.

**Y.C.** Conceptualization, Methodology, Supervision, Writing–Review & Editing.

**H.X.** Conceptualization, Methodology, Supervision, Resources, Funding acquisition, Writing–Review & Editing, Project administration.

**Y.Z.** Conceptualization, Methodology, Supervision, Funding acquisition, Writing–Original Draft, Writing–Review & Editing, Project administration.

All authors approved the final version of the manuscript. T.L., H.W., and Y.Z. accessed and verified the data underlying this study.

## Competing interests

The authors declare no competing interests.

## Notes

### Competing Interest Statement

The authors have declared no competing interest.

### Funding Statement

This study was funded by Shanghai Municipal Education Commission Program on AI for Transforming Academic Research Paradigms (24RGZNB05); Fudan University Medicine-Engineering Collaboration Program (yg-2025-general-07); The Medicine-X Innovation Team Incubation Program at the Children's Hospital of Fudan University (EKYX202413); The National Key Research and Development Program (2021YFC2500202); China Kidney Disease Big Data Collaboration Network Open Research Program (CRDS-004016).

### Author Declarations

The Ethics Committee of Children's Hospital of Fudan University gave ethical approval for this work (Approval No. 2022-05).

